# Social Class, Race/Ethnicity, and COVID-19 Mortality Among Working Age Adults in the United States

**DOI:** 10.1101/2021.11.23.21266759

**Authors:** Elizabeth B. Pathak, Janelle Menard, Rebecca B. Garcia, Jason L. Salemi

## Abstract

**Importance:** Substantial racial/ethnic disparities in COVID-19 mortality have been documented. Social class is a likely explanation of mortality disparities across and within racial/ethnic groups. This is the first U.S. study of social class and COVID-19 mortality in working age adults.

**Objectives:** To determine the joint effects of social class, race/ethnicity, and gender on the burden of COVID-19 mortality. A secondary objective was to determine whether differences in opportunities for remote work were correlated with COVID-19 death rates for sociodemographic groups.

**Design:** Annual mortality study which used a special government tabulation of 2020 COVID-19 related deaths stratified by decedents’ social class (educational attainment) and race/ethnicity.

**Setting:** United States in 2020.

**Participants:** COVID-19 decedents aged 25 to 64 years old (n=69,001).

**Exposures:** Social class (working class, some college, college graduate), race/ethnicity (Hispanic, Black, Asian, Indigenous, multiracial, and non-Hispanic white), and gender (women, men). Detailed census data on occupations held by adults in 2020 in each of the 36 sociodemographic groups studied were used to quantify the possibility of remote work for each group.

**Main Outcomes and Measures:** Age-adjusted COVID-19 death rates for 36 sociodemographic groups defined by social class, race/ethnicity, and gender. Disparities were quantified by relative risks and 95% confidence intervals. College graduates were the (low risk) referent group for all relative risk calculations.

**Results:** A higher proportion of Hispanics, Blacks, and Indigenous people were working class in 2020. COVID-19 mortality was five times higher in the working class vs. college graduates (72.2 vs. 14.6 deaths per 100,000, RR=4.94, 95% CI 4.82-5.05). The joint detriments of lower socioeconomic position, Hispanic ethnicity, and male gender resulted in a COVID-19 death rate which was over 27 times higher (178.0 vs. 6.5 deaths/100,000, RR=27.4, 95%CI 25.9-28.9) for working class Hispanic men vs. college graduate white women. In regression modeling, percent employed in never remote jobs explained 72% of the variance in COVID-19 death rates.

**Conclusions and Relevance:** SARS-CoV-2 infection control efforts should prioritize the working class (i.e. those with no college education), particularly those employed in “never remote” jobs with inflexible and unsafe working conditions (i.e. blue collar, service, and retail sales workers).

**KEY POINTS:** *Question:* Did COVID-19 mortality rates among non-elderly adults vary significantly by social class, race/ethnicity, and gender in 2020?

*Findings:* Among 69,001 COVID-19 decedents, age-adjusted COVID-19 deaths rates were 5 times higher in working class vs. college graduate adults 25-64 years old. Working class Hispanic, Black, and Indigenous men suffered the highest burden of COVID-19 mortality, while college graduate white women experienced the lowest death rate.

*Meaning:* COVID-19 mitigation efforts should prioritize the working class (i.e. those with no college education), particularly blue collar, service, and retail sales workers.

## BACKGROUND

COVID-19 is a viral infectious disease with a simple etiology (infection with the novel coronavirus SARS-CoV-2) and a complex clinical course which encompasses pathological derangement of multiple organ systems (e.g. respiratory,^1^ vascular,^2^ neurological,^3^ endocrine,^4^ and reproductive^5^); both an acute (days to weeks) and chronic (months to >1 year) clinical phase,^6,7^ and as yet unknown long-term clinical sequelae. Human-to-human transmission of SARS-CoV-2 occurs via exhalation of viral-laden aerosols by an infected person, suspension of these viral-laden aerosols in ambient air for extended periods of time, travel on expiratory plumes, and inhalation by susceptible persons at both near-field and far-field distances.^8-10^ Put simply, the social environments which can lead to SARS-CoV-2 infection are those in which people are breathing other people’s breath.^11,12^

Social class privilege creates the flexibility and space (quite literally) for the deployment of multiple strategies to reduce and prevent exposure to the highly infectious airborne novel coronavirus SARS-CoV-2. People in privileged social class positions live in larger homes with fewer people and in less densely populated neighborhoods (whether horizontally spacious in the suburbs or vertically spacious in metropolitan downtown areas), and rarely use public transportation. Additionally, the upper and professional classes have ready access to both high quality outpatient healthcare and the best tertiary care hospital centers.^13^ College education and related forms of social capital facilitate navigation of a complex healthcare system.^14^

Our study is the first national investigation of social class disparities in working age adult COVID-19 mortality, and it takes advantage of an *ad hoc* death certificate tabulation released by the U.S. National Center for Health Statistics (NCHS) in February 2021.^15,16^ These data permitted the calculation of age-adjusted COVID-19 mortality rates stratified *simultaneously* by social class, race/ethnicity, and gender. We take an explicitly anti-essentialist stance^17^ on the meaning of race and ethnicity^18^ in the epidemiology of COVID-19. Reductionist narratives of race and cultural, moral, and biological inferiority^19,20^ persist in public health and medicine when race is cited as an explanatory variable for negative health outcomes in the absence of social and historical contexts. We recognize race not as a genetic or physiological risk factor, but rather as a social construct^21^ that is embedded within a nexus of social oppression, exploitation, and conflict. This nexus amplifies exposure risks that result in higher burdens of morbidity and mortality among racial and ethnic minority populations.

The vast majority of working class adults are employed in blue collar, service, or retail sales jobs which require onsite attendance and prolonged close contact with others. In addition, working conditions vary by gender^22^ and race/ethnicity as well as social class.^23,24^ The most physically hazardous occupations are highly segregated by gender and performed largely by men (e.g. meatpacking). At the same time, under racialized capitalism, whites enjoy advantages of occupational status within social classes and even within narrowly defined job categories, compared with Hispanic, Black, and Indigenous workers.^24-26^ Moreover, elevated infection risks are amplified across multiple social environmental scales for working class adults,^27^ who may reside in poorly ventilated housing,^28^ commute in a crowded carpool, and labor in a crowded, poorly ventilated worksite.

We hypothesized that there were (1) significant social class disparities in working age adult mortality; (2) significant social class disparities in every racial/ethnic group; and (3) within-social class gender and racial/ethnic differences in opportunities for remote work that would be correlated with within- and between-class gender and racial/ethnic disparities in COVID-19 mortality.

## DATA AND METHODS

### Population at Risk

Our target population included adults aged 25 to 64 years who were U.S. residents during 2020. We included six racial/ethnic groups: whites, Hispanics, Blacks, Asians, Indigenous, and multi-race. The Indigenous group included American Indians, Alaska Natives, Native Hawaiians, and other Pacific Islanders, who were grouped together because of small numbers of deaths in some age-social class strata.

### Definition of Social Class

We used educational attainment as a proxy of social class, defined as follows: ***working class*** (no college), ***some college*** (including associate’s and other 2-year degrees), and ***college graduate*** (bachelor’s degree and higher).

### COVID-19 Deaths

***COVID-19 involved deaths*** included all deaths for which COVID-19 (ICD-10 code U07.1) was listed as the underlying or a contributing cause of death on the death certificate. We analyzed provisional death counts for 2020 stratifed by four sociodemographic variables: 1) <u>educational attainment</u> (no college, some college, college graduate); 2) <u>race and ethnicity</u> (white non-Hispanic, Hispanic, Black non-Hispanic, Asian non-Hispanic, American Indian/Alaska Native non-Hispanic, Native Hawaiian and other Pacific Islander non-Hispanic, more than one race non-Hispanic, unknown); 3) <u>gender</u> (male, female, unknown); and 4) <u>age group</u> (25-39 years, 40-54 years, 55-64 years).^15,16^

### Population Denominators

We used the 2020 Annual Social and Economic Supplement (ASEC) to the Current Population Survey (CPS) to calculate national population estimates stratified by educational attainment, race/ethnicity, gender, and age to exactly match the strata available in the COVID-19 death dataset.^29^

Public-use CPS datasets include statistical weights to calculate national population estimates from the household-based sample.^30^ We used special alternative weights that compensated for lower 2020 response rates in the CPS which were found to be differential by respondent income.^30,31^

### Death Rate Calculations

We first calculated age-specific death rates (deaths/population) for three age strata (25-39 years, 40-54 years, and 55-64 years) by social class for the following groups: (a) all adults combined; (b) men and women; (c) six racial/ethnic groups, and (d) 12 groups defined by both gender and race/ethnicity. Next, we verified that social class patterns of mortality were similar across age for all population groups. Then we calculated age-adjusted mortality rates for ages 25-64 combined, using the direct method with the U.S. 2020 population as the standard.

### Social Class Occupation Distributions

For the 36 sociodemographic groups aged 25 to 64 years (3 social class strata x 2 gender strata x 6 race/ethnicity strata), we used the 2020 CPS ASEC^29^ to identify the percent of adults with reported occupation in the following mutually-exclusive categories: (1) blue collar, (2) service, (3) retail sales, (4) health professionals, and (5) white collar (excluding health professionals and retail sales). Further details and specific examples of common job titles in each of these categories can be found in Table S1.

We rated each job title in the CPS on its potential for remote work (i.e. work from home). All blue collar, service, and retail sales jobs were classified as “never remote” jobs. All other jobs were classified as “sometimes remote” (health professionals) or “feasibly remote” (all other white collar jobs).

### Analytic Methods

We calculated social class rate ratios (RRs) of the age-adjusted death rates for the entire study population, by gender, by race/ethnicity, and finally by gender and race/ethnicity simultaneously. College graduates were the referent group for all comparisons. Then, we calculated disparity RRs that compared COVID-19 mortality in 35 sociodemographic groups with a single low-risk referent group (white women college graduates). Finally, we regressed the population-weighted log-transformed age-adjusted COVID-19 mortality rates against the percent of workers employed in never remote jobs for the 36 sociodemographic groups described above.

## RESULTS

There were 71,484 COVID-19 involved deaths among adults aged 25 to 64 years old during calendar year 2020 (Figure S1), as reported to NCHS by the end of February 2021. There were very few missing data; 2,483 deaths (3.5%) were excluded for missing race/ethnicity (0.5%) or social class (3.0%). The final analytic dataset included 96.5% of the total deaths (n=69,001) (Figure S1).

### Social Class Distribution of the Population at Risk

There were 168.4 million adults aged 25 to 64 years old in the U.S. in 2020. Figure 1 presents social class population pyramids for each of the 12 gender-race/ethnicity groups. In each pyramid, college graduates are represented in the top tier, those with some college in the middle tier, and the working class in the bottom tier. White men and women comprised approximately 60.2% of the total population at risk for working age COVID-19 mortality, and college graduates comprised the largest class among whites. Hispanics were predominantly working class. The working class also predominated among Black and Indigenous men.

**Figure 1.**
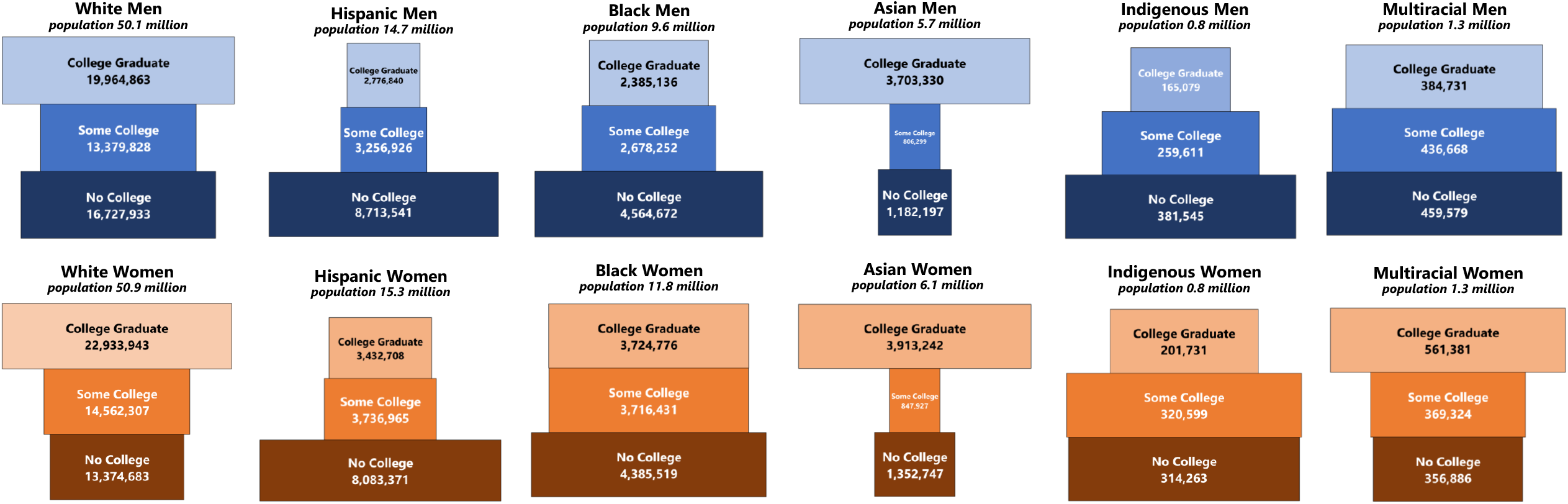
Social Class Population Pyramids,^*^ Adults 25-64 Years Old (n = 168.4 million) United States 2020. ^*^Note that the width of each bar is proportional to the size of the social class stratum within each race/ethnicity-gender group. Indigenous includes American Indians, Alaska Natives, Native Hawaiians, and other Pacific Islanders.

### Social Class and COVID-19 Mortality: Total Population

The age-adjusted COVID-19 mortality rate among college graduates aged 25-64 years was 14.6 deaths per 100,000 (Table 1). The death rate was twice as high among adults with some college but no 4-year degree (30.4 deaths/100,000; RR=2.08, 95% CI 2.02-2.14) and five times as high among working class adults (72.2 deaths/100,000; RR=4.94, 95% CI 4.82-5.05). The majority (68%, n=46,966) of COVID-19 decedents were working class, and only 12% (n=8,421) were college graduates.

**Table 1.** Social Class Disparities in Reported COVID-19 Mortality by Gender and Race/Ethnicity Among Adults 25-64 Year Old in the United States, 2020

| **Demographic Groups** | **Working Class** | **Some College** | **College Grads** |
| --- | --- | --- | --- |
| Population at risk | 59.9 million | 44.4 million | 64.1 million |
| **Total Study Population** |  |  |  |
| COVID-19 deaths | 46,966 | 13,614 | 8,421 |
| Age-adjusted mortality rate | 72.2/100,000 | 30.4/100,000 | 14.6/100,000 |
| Social class rate ratio (95% CI) | 4.94 (4.82-5.05) | 2.08 (2.02-2.14) | 1.0 (referent) |
| **By Reported Gender** |  |  |  |
| **Women** |  |  |  |
| COVID-19 deaths | 15,708 | 5,535 | 3,039 |
| Age-adjusted mortality rate | 50.4/100,000 | 22.8/100,000 | 10.0/100,000 |
| Social class rate ratio (95% CI) | 5.06 (4.87-5.26) | 2.29 (2.19-2.39) | 1.0 (referent) |
| **Men** |  |  |  |
| COVID-19 deaths | 31,258 | 8,079 | 5,382 |
| Age-adjusted mortality rate | 92.1/100,000 | 39.5/100,000 | 19.8/100,000 |
| Social class rate ratio (95% CI) | 4.65 (4.52-4.79) | 1.99 (1.93-2.06) | 1.0 (referent) |
| **By Reported Race/Ethnicity** |  |  |  |
| **White, non-Hispanic** |  |  |  |
| COVID-19 deaths | 14,587 | 5,344 | 3,746 |
| Age-adjusted mortality rate | 40.6/100,000 | 17.8/100,000 | 9.3/100,000 |
| Social class rate ratio (95% CI) | 4.37 (4.21-4.53) | 1.92 (1.84-2.00) | 1.0 (referent) |
| **Hispanic** |  |  |  |
| COVID-19 deaths | 19,174 | 3,173 | 1,540 |
| Age-adjusted mortality rate | 125.0/100,000 | 57.0/100,000 | 32.9/100,000 |
| Social class rate ratio (95% CI) | 3.80 (3.61-4.00) | 1.73 (1.63-1.84) | 1.0 (referent) |
| **Black, non-Hispanic** |  |  |  |
| COVID-19 deaths | 10,544 | 3,912 | 1,989 |
| Age-adjusted mortality rate | 105.9/100,000 | 59.0/100,000 | 33.8/100,000 |
| Social class rate ratio (95% CI) | 3.14 (2.99-3.29) | 1.75 (1.66-1.84) | 1.0 (referent) |
| **Asian, non-Hispanic** |  |  |  |
| COVID-19 deaths | 1,149 | 497 | 955 |
| Age-adjusted mortality rate | 38.5/100,000 | 32.1/100,000 | 17.7/100,000 |
| Social class rate ratio (95% CI) | 2.18 (2.00-2.38) | 1.82 (1.63-2.03) | 1.0 (referent) |
| **Indigenous, non-Hispanic** |  |  |  |
| COVID-19 deaths | 1,353 | 602 | 137 |
| Age-adjusted mortality rate | 182.1/100,000 | 113.4/100,000 | 37.2/100,000 |
| Social class rate ratio (95% CI) | 4.90 (4.11-5.84) | 3.05 (2.53-3.67) | 1.0 (referent) |
| **Multirace/Other, non-Hispanic** |  |  |  |
| COVID-19 deaths | 159 | 86 | 54 |
| Age-adjusted mortality rate | 20.0/100,000 | 12.9/100,000 | 8.7/100,000 |
| Social class rate ratio (95% CI) | 2.32 (1.70-3.15) | 1.49 (1.06-2.10) | 1.0 (referent) |

### Social Class and COVID-19 Mortality by Gender

Women experienced lower COVID-19 death rates than men (college grad women 10.0 deaths/100,000 vs. 19.8 deaths/100,000 in college grad men), but a slightly higher social class disparity (RR=5.06, 95% CI 4.87-5.26 in women vs. RR=4.65, 95% CI 4.52-4.79 in men) for the working class vs. college graduates. Numerically, both the age-adjusted death rate (92.1/100,000) and the number of deaths (n=31,258) were highest for working class men (Table 1).

### Social Class and COVID-19 Mortality by Race and Hispanic Ethnicity

In all six racial/ethnic groups, there was a monotonic association between social class and COVID-19 mortality, with the lowest age-adjusted death rates among college graduates, and the highest rates among the working class (Table 1). Social class disparity RRs ranged from 2.18 (95% CI 2.00-2.38) among Asians to RR=4.90 (95% CI 4.11-5.84) among Indigenous adults. Within each social class, death rates were highest for Indigenous, Hispanic, and Black adults, and lowest for multiracial, Asian, and white adults.

### Disparities in COVID-19 Mortality: Independent and Joint Effects of Social Class, Gender, and Race/Ethnicity

The independent effects of social class, gender, and race/ethnicity on COVID-19 mortality are evident in Figure 2 for Hispanics, Blacks, and whites, who together comprised 90.5% of the total population at risk. Across all six groups defined by gender and race/ethnicity, there was a strong and statistically significant association of social class with age-adjusted COVID-19 mortality (see Table S2 for all RRs and 95%CI). Similarly, across all nine groups defined by social class and race/ethnicity, age-adjusted death rates were always higher for men than for women. However, there was effect modification by gender when stratifying by social class. Among men, the highest death rates were suffered by Hispanics in each social class. But among women, Blacks suffered the highest death rates in each social class.

**Figure 2.**
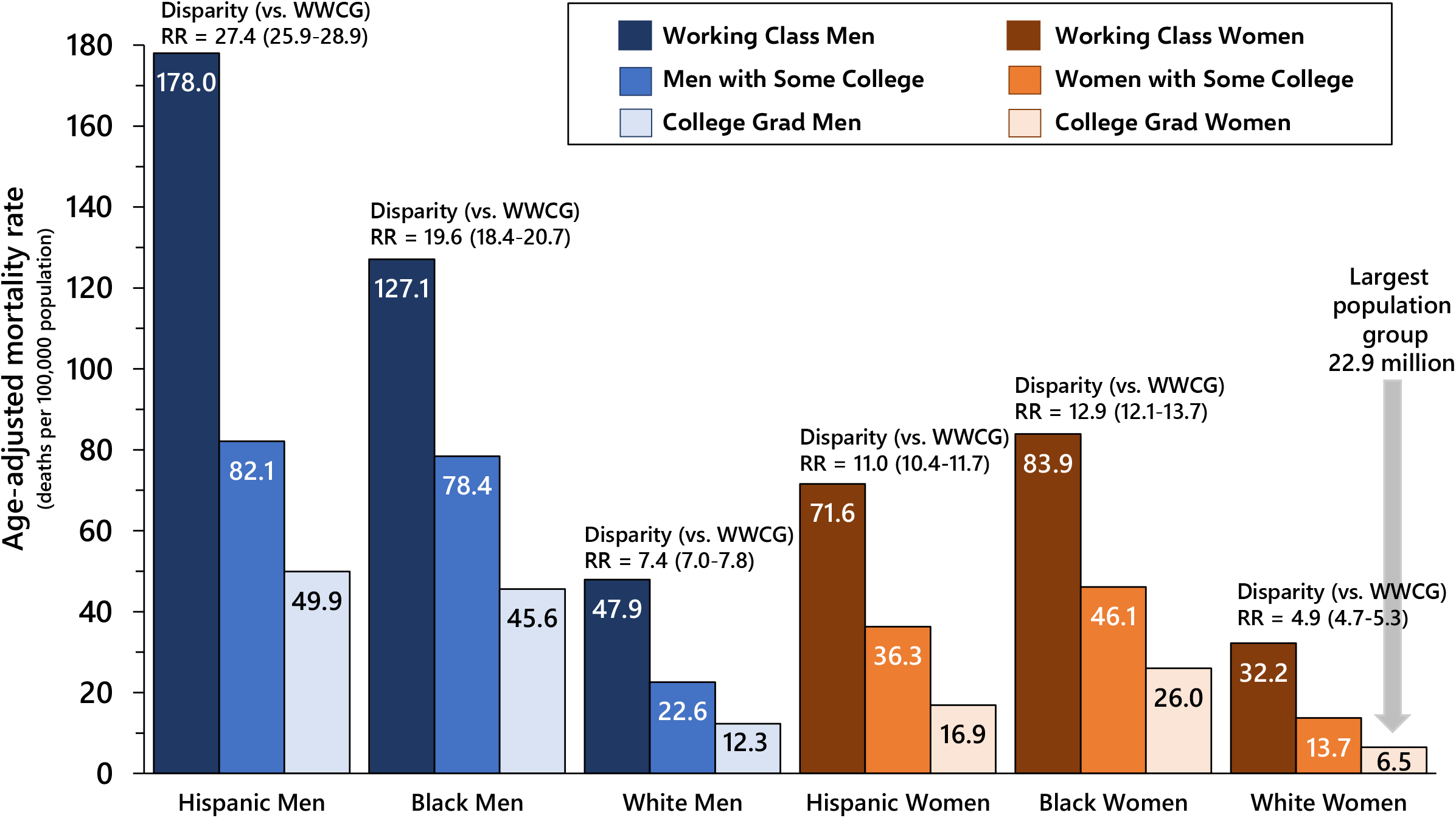
COVID-19 Death Rates and Social Class-Gender-Race/Ethnicity Disparity Rate Ratios,^**^ with White Women College Graduates (WWCG) as the Referent Group Adults 25-64 Years Old, United States 1-Jan-2020 to 31-Dec-2020. ^**^ The disparity rate ratios (RR) are calculated separately for each socio-demographic group and compare age-adjusted COVID-19 death rates, with white women college graduates (WWCG) as the referent group. Selected results are presented for the 3 largest population groups (Whites, Hispanics, and Blacks). Full results, including for Asians, Indigenous adults (American Indians, Alaska Natives, Native Hawaiians, and other Pacific Islanders) and multiracial adults are shown in Table S2.

Finally, disparity RRs which capture the joint effects of social class, gender, and race/ethnicity on working age COVID-19 mortality confirm that white women college graduates were at lowest risk for COVID-19 mortality (6.5 deaths/100,000). The joint detriments of working class socioeconomic position, Hispanic ethnicity, and male gender resulted in a COVID-19 age-adjusted death rate which was over 27 times higher (178.0 deaths/100,000, RR=27.4, 95%CI 25.9-28.9) (Figure 2). While in all social classes Hispanic and Black women experienced lower death rates than Hispanic and Black men, respectively, they suffered higher death rates than white men across social classes. Full results for all 36 sociodemographic groups are available in Table S2.

### Low-Level Jobs and Never Remote Work by Social Class, Gender, and Race/Ethnicity

As expected, the majority of employed college graduates had white collar jobs, and those with some college were employed in a mixture of blue collar, service, retail sales, and white collar jobs, with no category in the majority (Figure 3). Conversely, the majority of working class adults were employed in the lowest-level jobs (blue collar, service, and retail sales) with no potential for remote work. However, majority employment in low-level jobs varied from 51.1% of working class white women to 85.9% of working class non-white men. Across social classes, non-whites were more likely to be employed in service jobs than whites, and men were much more likely to be employed in blue collar jobs than women.

**Figure 3.**
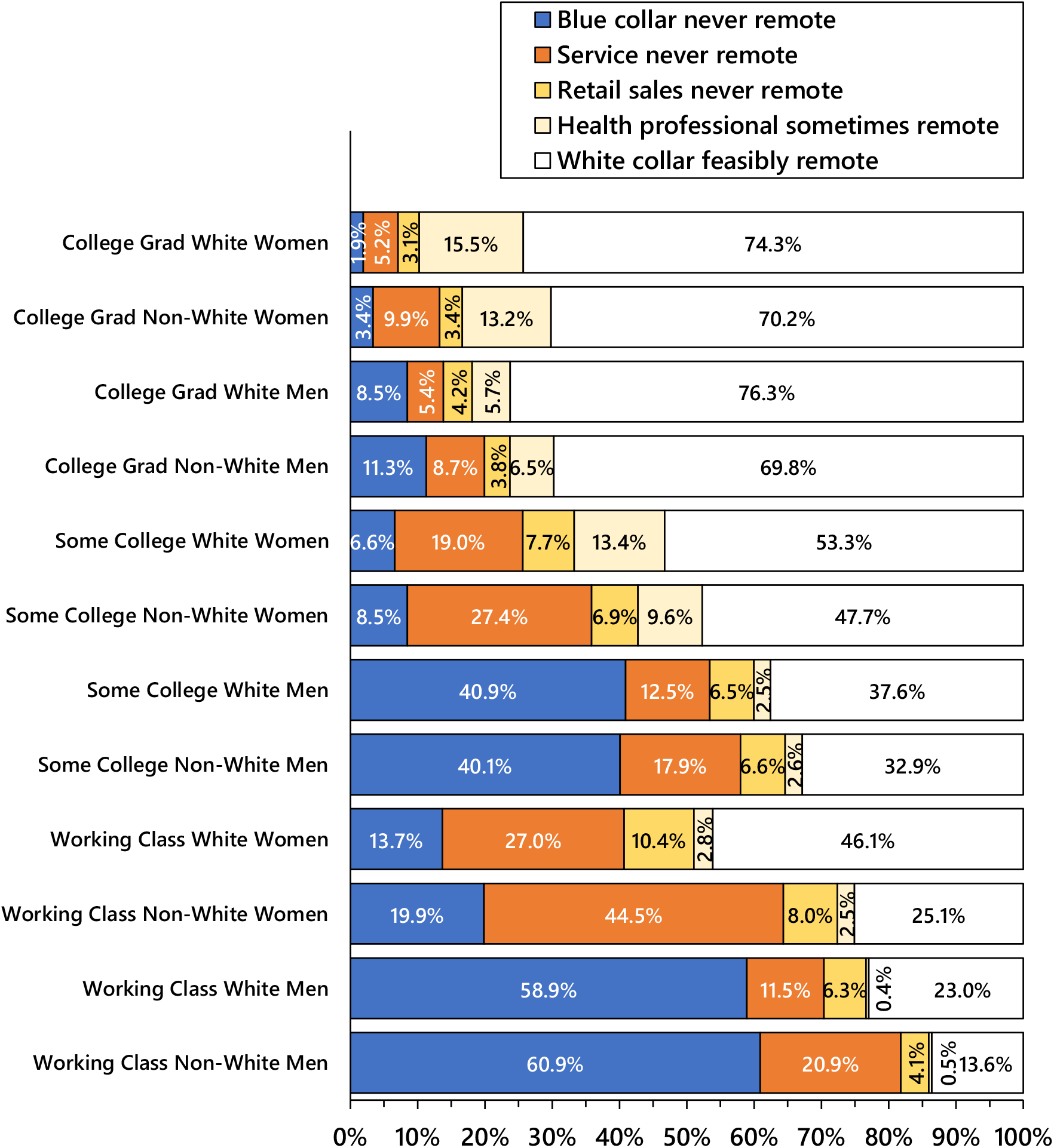
Remote Work Occupations by Social Class, Gender, and Race/Ethnicity, Adults 25-64 Years Old, United States 2020. Non-White includes Hispanic, Black, Asian, Indigenous, and Multiracial adults. Service is comprised of healthcare support, protective service, food service, housekeeping, building and grounds, and personal care service workers. Health professionals include registered nurses and licensed practical nurses. Blue collar includes transportation workers, including airline pilots and flight attendants. White collar feasibly remote is comprised of managers, professionals, technical workers, non-retail sales workers, and office support and administrative workers.

A population-weighted regression of the natural log-transformed age-adjusted 2020 COVID-19 death rates revealed a good fit of an exponential model in which the percent of adults employed in never remote jobs during 2020 explained 72% of the variance in the age-adjusted death rates across the 36 population groups defined by social class, gender, and race/ethnicity (Figure 4).

**Figure 4.**
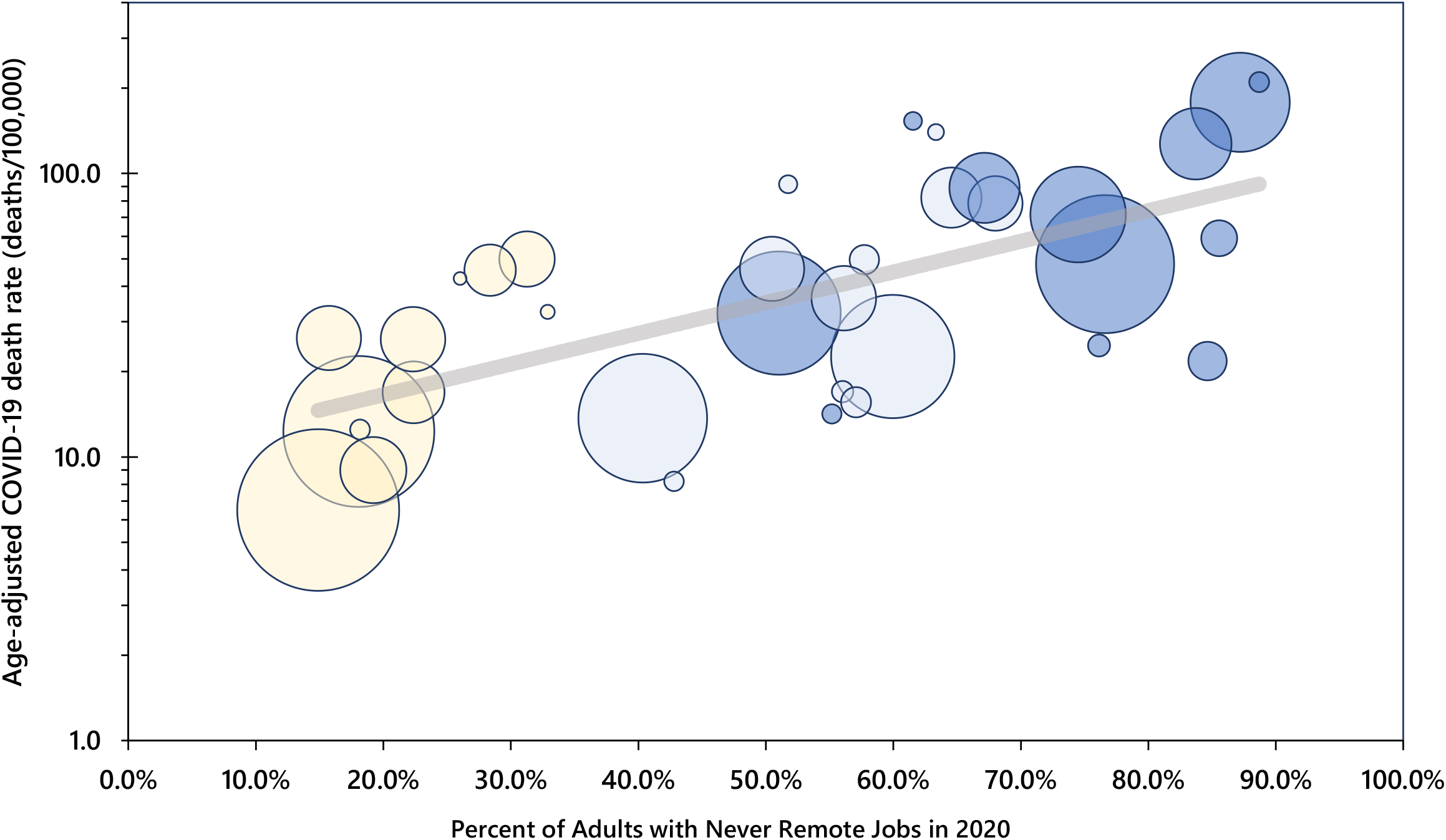
COVID-19 Mortality by Never Remote (Blue Collar/Service/Retail) Jobs Adults 25 to 64 Years Old, United States 2020. Each bubble represents a social class, gender, racial/ethnic group, with bubble size proportional to population size. Dark blue = working class, light blue = some college, yellow = college grads..

## DISCUSSION

For individuals, the accumulation of economic and social capital gives rise to a privileged social class position, which grants power relative to others in society.^32^ Class power manifests itself through control over economic resources (including labor and the means of production) and in a high social status that opens access to less tangible privileges via well-resourced social networks.^33^

Most importantly, people with social class power and privilege retain a far greater degree of discretionary control over their professions, work lives, and daily schedules than the working class. For many, a college degree and professional status provides a measure of autonomy and flexibility in meeting job requirements.^34^ In contrast, the working class (in blue collar, service, and retail sales occupations) are subjected to authoritarian control^35^ and inflexible requirements of work.^24,34,36,37^ Moreover, the worksites in which the working class perform their wage labor often are replete with physical, chemical, and biologic hazards which directly and negatively impact workers’ health and well-being.^34,38-40^

In the United States, social class is an intrinsically racialized set of economic and social status relationships.^41-43^ The legacies of colonialism, slavery, and other forms of structural racism shape local labor markets, housing opportunities, and other material aspects of workers’ lives.^23,44^ Consequently, a given level of educational attainment usually provides fewer economic benefits to Blacks and other minorities.^23,44^

Our results support the hypothesis that hazardous material conditions of work were a primary driver of joint social class and racial/ethnic disparities in COVID-19 mortality. During the first year of the COVID-19 pandemic in the United States, working class adults aged 25-64 years old were five times as likely as college graduates to die from COVID-19, and adults with some college but no four-year degree were twice as likely as college graduates to die. White college graduates aged 25 to 64 years were largely shielded from COVID-19 mortality during the first year of the pandemic. They comprised more than one-quarter of the study population, but accounted for only 5% of the COVID-19 deaths. White women college graduates, the numerically largest population group (n=22.9 million), accounted for only 2% of working age COVID-19 decedents. In contrast, Hispanic and Black working class men comprised only 8% of the 25-64 year old population, but they were 29% of the premature COVID-19 decedents. Non-white working class men were most likely to be employed in never remote occupations (i.e. blue collar, service, and retail sales) compared with every other sociodemographic group.

Our results are consistent with those of a smaller study of excess mortality by occupation in California during March – October 2020,^45^ and with a small study of worksite COVID-19 transmission in Asian countries which found the most commonly affected occupations were healthcare, drivers, sales, cleaners, and public safety.^46^ A major report on social inequalities in COVID-19 in the United Kingdom found social class patterns of COVID-19 mortality that were very similar to what we observed for the U.S.^28^ However, the magnitude of the social class mortality disparities was much lower in the U.K.

### COVID-19 Case Fatality

Axiomatically, mortality rates (deaths/population) are a function of two underlying phenomena: the incidence of disease in a specified population (cases/population) and the case fatality rate (deaths/cases) of the disease. We hypothesize that disparities in both case fatality and incidence have contributed to the strong and highly significant mortality disparities observed in our study. Access to high quality evidence-based medical care is not universal in the U.S.^47^ Barriers to accessing timely and appropriate COVID-19 medical care include lack of health insurance, inadequate health insurance (e.g. high deductible/co-pay plans), lack of or inadequate paid sick leave,^48^ geographic location, transportation access/costs/timeliness, lack of respite dependent care, threat of job loss, immigration status, racism and discrimination, and distrust of healthcare and government institutions.^49^ An analysis of place of death of U.S. COVID-19 decedents found that 22% of 30-49 year olds and 14% of 50-64 year olds died either outside a hospital or in the emergency department (OH/ED).^50^ Minimizing COVID-19 case fatality requires that individuals have access to timely diagnosis and high-quality hospital medical care before they become critically ill.

### Study Limitations and Public Health Data Gaps

It is likely that COVID-19 deaths in the U.S. have been undercounted (i.e., cause of death has been misclassified), and this misclassification is likely to be differential by social class, resulting in a bias toward the null in our estimates of social class disparities. Misclassification occurs when there is insufficient medical information available at the time of death. Lack of access to medical care and out-of-hospital mortality can result in the use of non-specific cause of death coding on death certificates. We have previously shown that the percent of all non-injury deaths coded to “symptoms, signs, and ill-defined conditions” increased from 2019 to 2020 among working age adults.^50^

A simple step toward improving COVID-19 surveillance data, which could be implemented immediately across a wide range of data systems, is to add one yes/no question to all individual adult patient encounter medical records: “Has this person completed one or more years of college?” A “no” response on this single data item would identify the working class. A follow-up question for those who replied “yes” (“Does this person have a 4-year college degree?”) would easily identify the three social classes analyzed in this study.

## Conclusion

The most urgent implication of our study points to immediate actions needed to protect blue collar, service, and retail sales workers from infection with the SARS-CoV-2 virus. Expert recommendations include strengthening federal and state labor laws,^51^ empowering OSHA,^40^ adopting the Total Worker Health Framework,^52^ and direct actions for unions to organize for greater protections for worker safety.^39^

## Data Availability

All data analyzed in this study are publicly available online.

---

**Figure S1.**
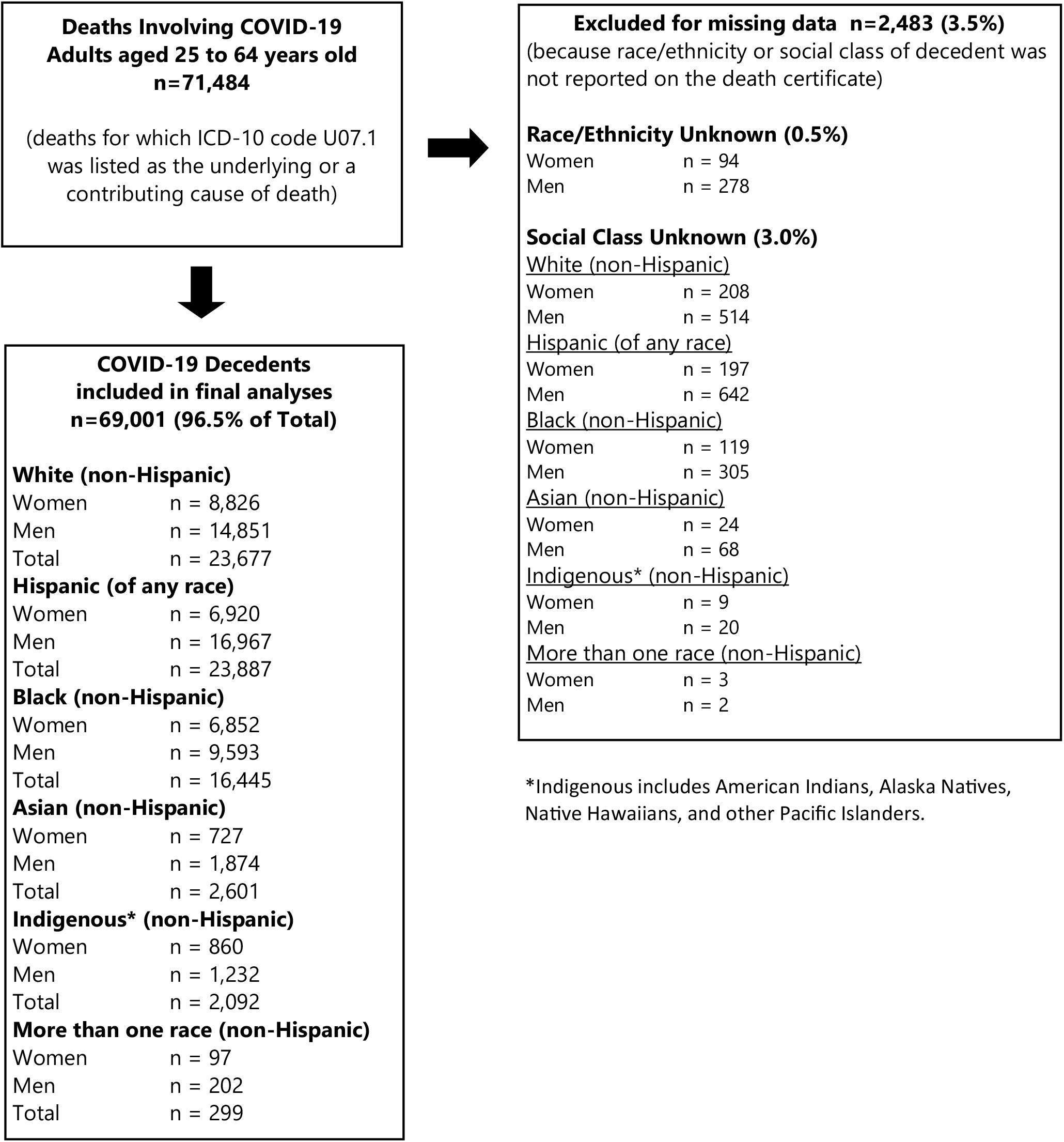
Study Inclusion of Deaths Involving COVID-19 Adults Aged 25 to 64 Years Old, United States, 1-Jan-2020 to 31-Dec-2020

**Table S1.** Top 5 Occupations of Adults 25-64 years old in 2020, by Social Class, Gender, and Race/Ethnicity. **Blue =** Blue collar, never remote **Orange =** Service, never remote **Gold =** Retail sales, never remote **Yellow =** Health professionals, rarely remote **White =** White collar, usually remote

| Six Most Populous Groups | Working class | Some college | College grads |
| --- | --- | --- | --- |
| **White men** | 1. Driver/sales workers, and truck drivers<br>2. Construction laborers<br>3. Managers, other<br>4. Carpenters<br>5. Laborers, hand material movers | 1. Managers, other<br>2. Driver/sales workers, and truck drivers<br>3. Retail supervisors<br>4. Electricians<br>5. Retail salespersons | 1. Managers, other<br>2. Software developers<br>3. Chief executives, legislators<br>4. Lawyers, judges, magistrates<br>5. Elementary, middle school teachers |
| **Hispanic men** | 1. Construction laborers<br>2. Driver/sales workers, and truck drivers<br>3. Landscaping, grounds maintenance workers<br>4. Carpenters<br>5. Laborers, hand material movers | 1. Driver/sales workers, and truck drivers<br>2. Construction laborers<br>3. Retail salespersons<br>4. Managers, other<br>5. Retail supervisors | 1. Managers, other<br>2. Elementary, middle school teachers<br>3. Software developers<br>4. Accountants and auditors<br>5. Computer occupations, all other |
| **Black men** | 1. Driver/sales workers, and truck drivers<br>2. Laborers, hand material movers<br>3. Janitors and building cleaners<br>4. Cooks<br>5. Construction laborers | 1. Driver/sales workers, and truck drivers<br>2. Laborers, hand material movers<br>3. Retail salespersons<br>4. Security guards, gaming surveillance officers<br>5. Stockers and order fillers | 1. Managers, other<br>2. Elementary, middle school teachers<br>3. Software developers<br>4. Driver/sales workers, truck drivers<br>5. Accountants and auditors |
| **White women** | 1. Secretaries and administrative assistants<br>2. Retail supervisors<br>3. Cashiers<br>4. Bookkeeping and accounting clerks<br>5. Customer service representatives | 1. Registered nurses<br>2. Secretaries and administrative assistants<br>3. Retail supervisors<br>4. Bookkeeping and accounting clerks<br>5. Managers, other | 1. Elementary/middle school teachers<br>2. Registered nurses<br>3. Managers, other<br>4. Accountants and auditors<br>5. Secondary school teachers |
| **Hispanic women** | 1. Maids and housekeepers<br>2. Janitors and building cleaners<br>3. Cooks<br>4. Personal care aides<br>5. Cashiers | 1. Secretaries, administrative assistants<br>2. Cashiers<br>3. Office clerks, general<br>4. Customer service representatives<br>5. Bookkeeping and accounting clerks | 1. Elementary/middle school teachers<br>2. Registered nurses<br>3. Managers, other<br>4. Education administrators<br>5. Secretaries, administrative assistants |
| **Black women** | 1. Nursing assistants<br>2. Maids and housekeepers<br>3. Personal care aides<br>4. Cashiers<br>5. Home health aides | 1. Nursing assistants<br>2. Customer service representatives<br>3. Registered nurses<br>4. Licensed practical nurses<br>5. Secretaries, administrative assistants | 1. Registered nurses<br>2. Elementary/middle school teachers<br>3. Managers, other<br>4. Social workers<br>5. Accountants and auditors |

Blue collar included construction, mining, farming, forestry, installation, maintenance, and repair jobs, fabrication and production (i.e., manufacturing) jobs, transportation, warehousing, material moving, and general manual labor jobs. Service jobs included housekeeping, janitorial, groundskeeping, food service, protective services, personal care, childcare, and healthcare service. White collar jobs included all managerial, professional (except health professionals), technical, sales (except retail sales), and administrative and office support occupations.

**Table S2.**
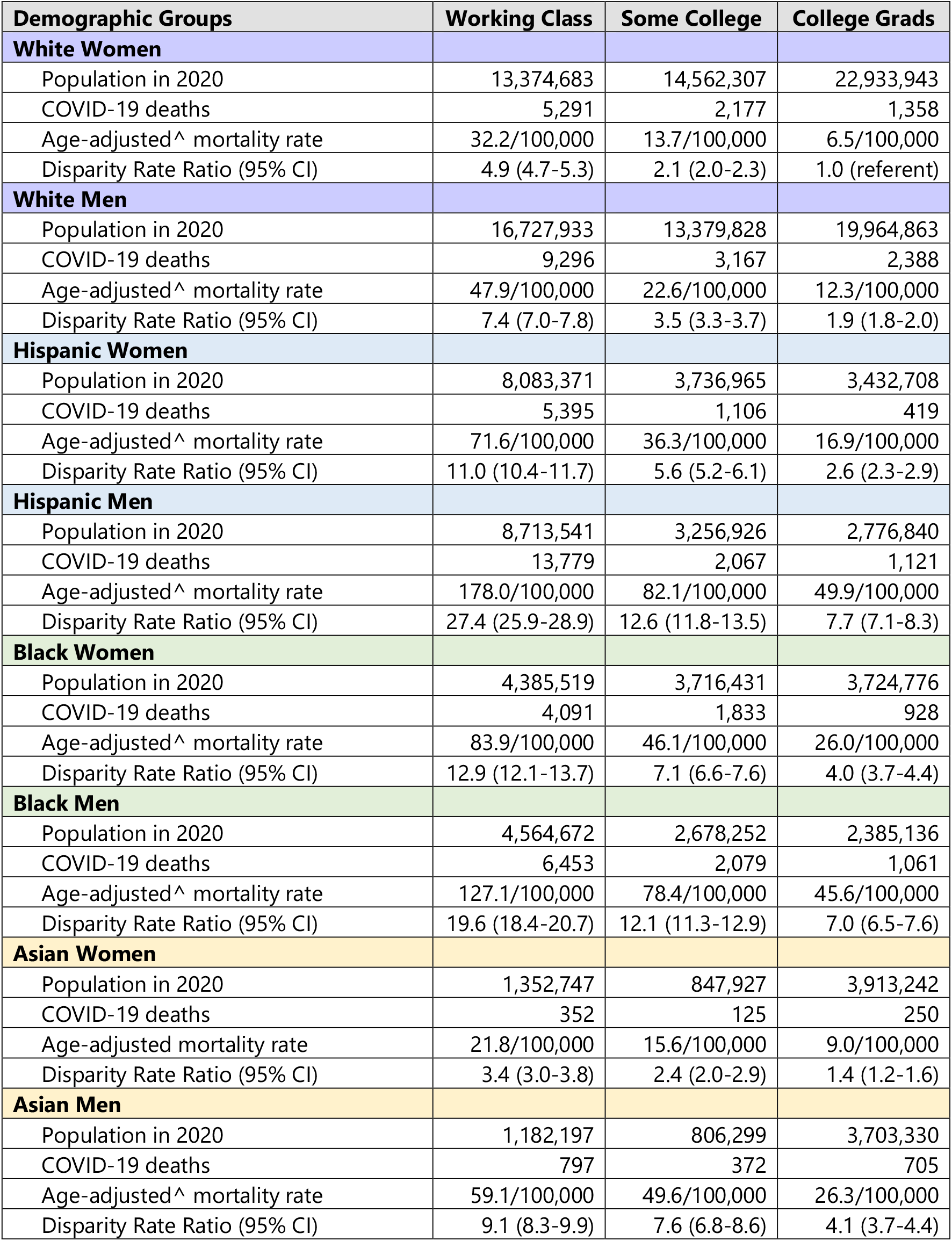
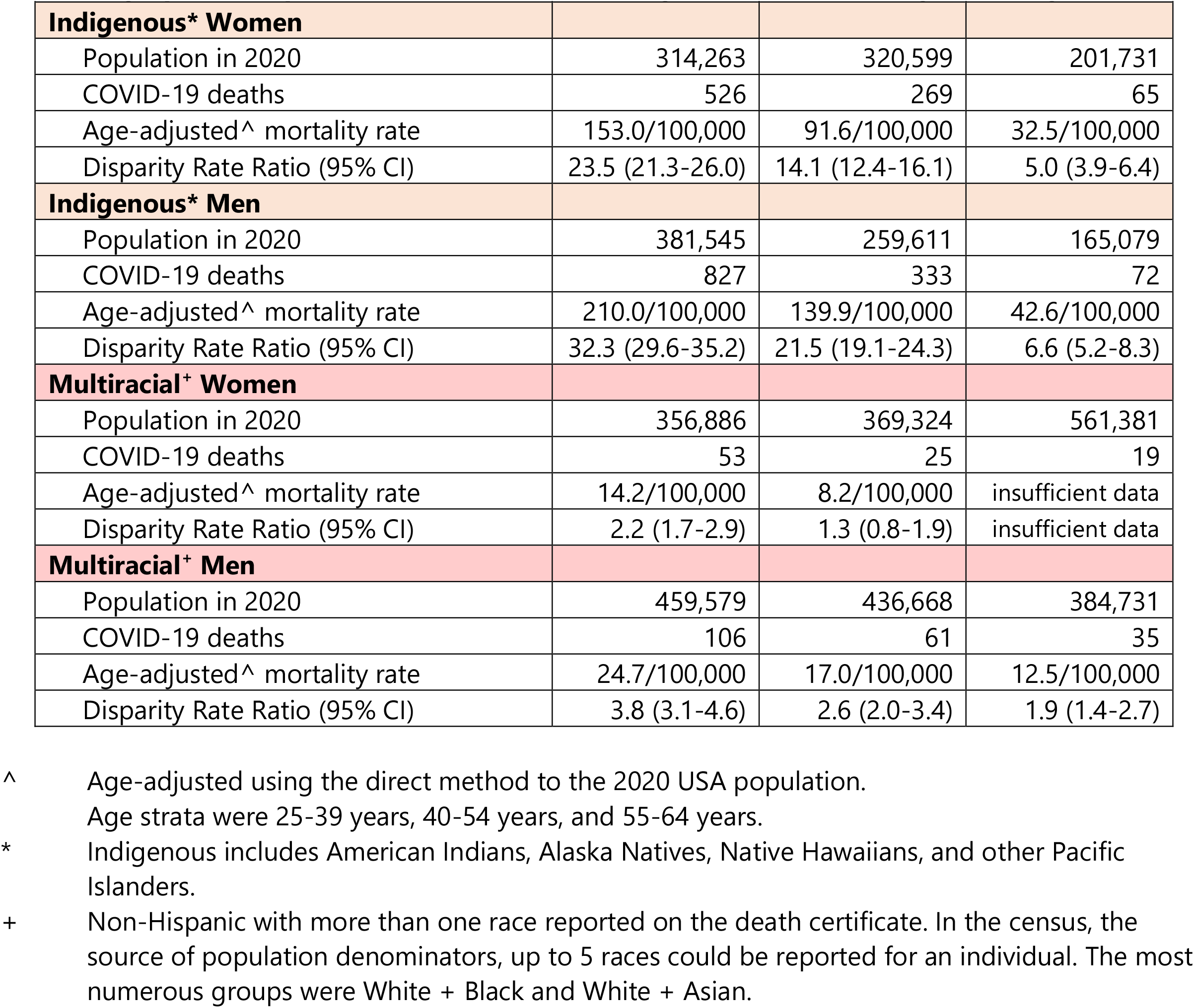
Disparities in Reported COVID-19 Mortality by Social Class, Race/Ethnicity, and Gender Among Adults 25-64 Year Old in the United States, 1-Jan-2020 to 31-Dec-2020 (White Women College Graduates are the Referent Group for all Disparity Rate Ratios)

